# The effect of High Definition Transcranial Direct Current Stimulation (HD-tDCS) on pain in Somatic Symptom Disorder with predominant pain (SSD-P): a randomized single-blind sham controlled cross-over study

**DOI:** 10.1101/2023.08.01.23293484

**Authors:** Sanjana Kathiravan, Abhishek Ghosh, Shubh Mohan Singh

**Author notes:** Corresponding author name-Shubh Mohan Singh Number-9417249900 Email address- Professor, Department of Psychiatry, Postgraduate Institute of Medical Education and Research Chandigarh-160012, India.

## Abstract

**Background:** Somatic Symptom Disorder with predominant pain (SSD-P) is a common disorder with unsatisfactory treatment options. Transcranial Direct Current Stimulation (tDCS) is useful in chronic pain conditions.

**Objective:** To assess the effect of two within session repeated High Definition tDCS (HD-tDCS) over left motor cortex on pain and other associated variables in SSD-P.

**Methods:** Institute Ethics Committee approval was sought and the study was registered in the Clinical Trials Registry of India. Single-blind, sham controlled, cross-over study design was employed. Thirty right handed patients with DSM-5 diagnosis of SSD-P, aged 18-60 years, receiving stable treatment were enrolled by consecutive sampling. After simple randomization, two short interval repeated sessions (2mA, 20 minutes each) of either active or sham HD-tDCS was administered followed by a wash out period of seven days and cross over. Assessments were done at baseline, week one and two. Analysis was done using SPSS 21.

**Results:** Participants were comparable on all baseline parameters. ANCOVA showed no order×group interaction. On RM-ANOVA all participants irrespective of entering active or sham arm showed statistically significant difference (Cohen’s d >0.8) in pain and associated parameters like depression, anxiety, pain related interference, burden and disability at week two (p< 0.01). But at week one, this was observed only in active arm. Transient mild local side effects like burning, pain and itching were noted, with no cognitive side effects.

**Conclusion:** This was the first study of HD-tDCS on SSD-P. Single session repeated protocol can be effective in pain reduction with sustained effects up to one week.

## 1. Introduction

Pain is defined as ‘an unpleasant sensory and emotional experience associated with, or resembling that associated with, actual or potential tissue damage’ as per the International Association for the Study of Pain(Raja et al., 2020). Somatic symptom disorder (SSD) as per the Diagnostic and Statistical Manual of Mental Disorders, Fifth Edition (DSM-5) consists of somatic symptoms that are distressing or result in significant disruption in daily life persisting beyond 6 months(American Psychiatric Association, 2013). Somatic symptom disorder with predominant pain (SSD-P), a specifier for SSD has replaced the previous DSM-IV pain disorder with removal of the requirement that symptoms be medically unexplained(American Psychiatric Association, 2013; Katz et al., 2015). Although the exact estimate is not known, prevalence of SSD-P in the population stands at about 4% and associated with significant burden(Dimsdale et al., 2013; Henningsen, 2018). Cochrane review of pharmacological treatment options showed low quality evidence for antidepressants or natural products in SSD-P when compared to placebo(Kleinstäuber et al., 2014). Apart from cognitive behavioural therapy (CBT), all other forms of psychological therapies for SSD-P showed small effect sizes(van Dessel et al., 2014). However, SSD-P is considered “difficult to treat” as existing treatment modalities are associated with challenges(Henningsen, 2018).

Transcranial Direct Current Stimulation (tDCS) has been utilized in the treatment of various neurological and psychological conditions including chronic pain(Lefaucheur et al., 2017; Tortella et al., 2015). There is a lack of consensus in existing protocols, with most studies utilizing conventional tDCS using variable and multiple sessions(Lefaucheur, 2016; Lefaucheur et al., 2017). However, the effect of a single tDCS session on chronic pain conditions has been assessed in a few studies, showing significant pain relief when compared to sham stimulation(Bolognini et al., 2013; Fenton et al., 2009; Mendonca et al., 2011; Ngernyam et al., 2015). Moreover, very few studies have studied the effect of High Definition tDCS (HD-tDCS) on chronic pain(Donnell et al., 2015; Villamar et al., 2013).

HD-tDCS has not been studies in SSD-P. We followed a novel protocol of two sessions of HD-tDCS delivered within a 30 minute interval. This is a modification of single session protocols, wherein it has been seen that when two sessions are delivered within a short interval (within session repeated tDCS), the effects last longer due to increased excitability induced(Bastani and Jaberzadeh, 2014). The current study aims to investigate the effect of two within session repeated HD-tDCS on pain and other associated variables like burden, interference and disability in SSD-P.

## 2. Methods

### 2.1 Study design and setting

A single-blind, sham controlled, cross-over study design was employed compared to the traditional parallel-group design as within patient variation is less. The treatment periods were separated by a wash out period of 7 days. The study sample was recruited from November 2020 to May 2021 from the outpatient clinic of the Department of Psychiatry and Neurology of our tertiary care hospital after obtaining written informed consent. The study included patients presenting with chronic pain who fulfilled DSM-5 diagnosis of SSD-P. Written informed consent was obtained from all the study participants. Approval was sought from the Ethics Committees of the Institute in which this study was conducted (INT/IEC/2020/SPL-481). The study was registered in the Clinical Trials Registry of India (CTRI/2020/10/028752). No change to protocol was made after initiation of study.

### 2.2 Inclusion criteria

Right handed patients with diagnosis of SSD-P as per DSM-5, aged 18 to 60 years of either gender, providing written informed consent, and already receiving any form of stable treatment, pharmacological or non-pharmacological (with no change since the past three months) were included. It was explained to the patients that no change will be made to the treatment that the patient was already receiving (tDCS sessions would be an add-on treatment).

### 2.3 Exclusion criteria

Patients who did not give consent for study, those with diagnosed Parkinson’s disease, organic brain syndrome, intellectual disability or dementia, head injury, uncontrolled epilepsy, encephalopathy due to any cause or tobacco dependence, presence of other cross-sectional or lifetime comorbid psychiatric disorders (as determined by using MINI, except cross-sectional diagnosis of an anxiety or depressive disorder) and pregnant females were excluded from the study.

In view of the cross over design and as per the available data on the analgesic benefit of tDCS(Janice Jimenez-Torres et al., 2017), a total of 36 participants (18 per group randomized to the active tDCS or the sham tDCS conditions) was needed to detect an effect (alpha = .05, effect size f = .25, correlation among repeated measures = .5) with adequate power (1 – β = 0.95). But due to the COVID-19 pandemic and lockdown, we could recruit a sample size of 30 patients only and the corrected sample size was approved by the Institute.

### 2.4 Assessments

Baseline assessment was done at the point of entry into the study. It consisted of sociodemographic and clinical profile sheet, assessment of somatic symptoms using the Patient Health Questionnaire Physical Symptoms (PHQ-15) and Somatic Symptoms Experiences Questionnaire (SSEQ), and handedness by Edinburgh Handedness Inventory. Clinical assessment was done immediately before the commencement of the HD-tDCS session. It included assessment of pain on Numerical Rating Scale (NRS) and Brief Pain Inventory-Hindi version (BPI-H), assessment of depressive and anxiety symptoms on Patient Health Questionnaire (PHQ-9) and Generalized Anxiety Disorder 7 Scale (GAD 7), assessment of burden and disability due to pain on Somatic Symptom Scale–8 (SSS-8) and Pain Disability Index 7 (PDI), cognitive functions on Mini Mental State Assessment (MMSE). Further, Clinical Global Impression (CGI) and Patient global impression of change (PGIC) scales were applied to assess the overall severity and improvement with intervention. This was repeated at three time points-at baseline (week zero), end of week one and week two.

Immediate assessment was done to assess the perception of active or sham stimulation received and evaluation of side effects using tDCS side effect checklist. Figure 1 shows the flowchart of the proposed study.

**Figure 1.**
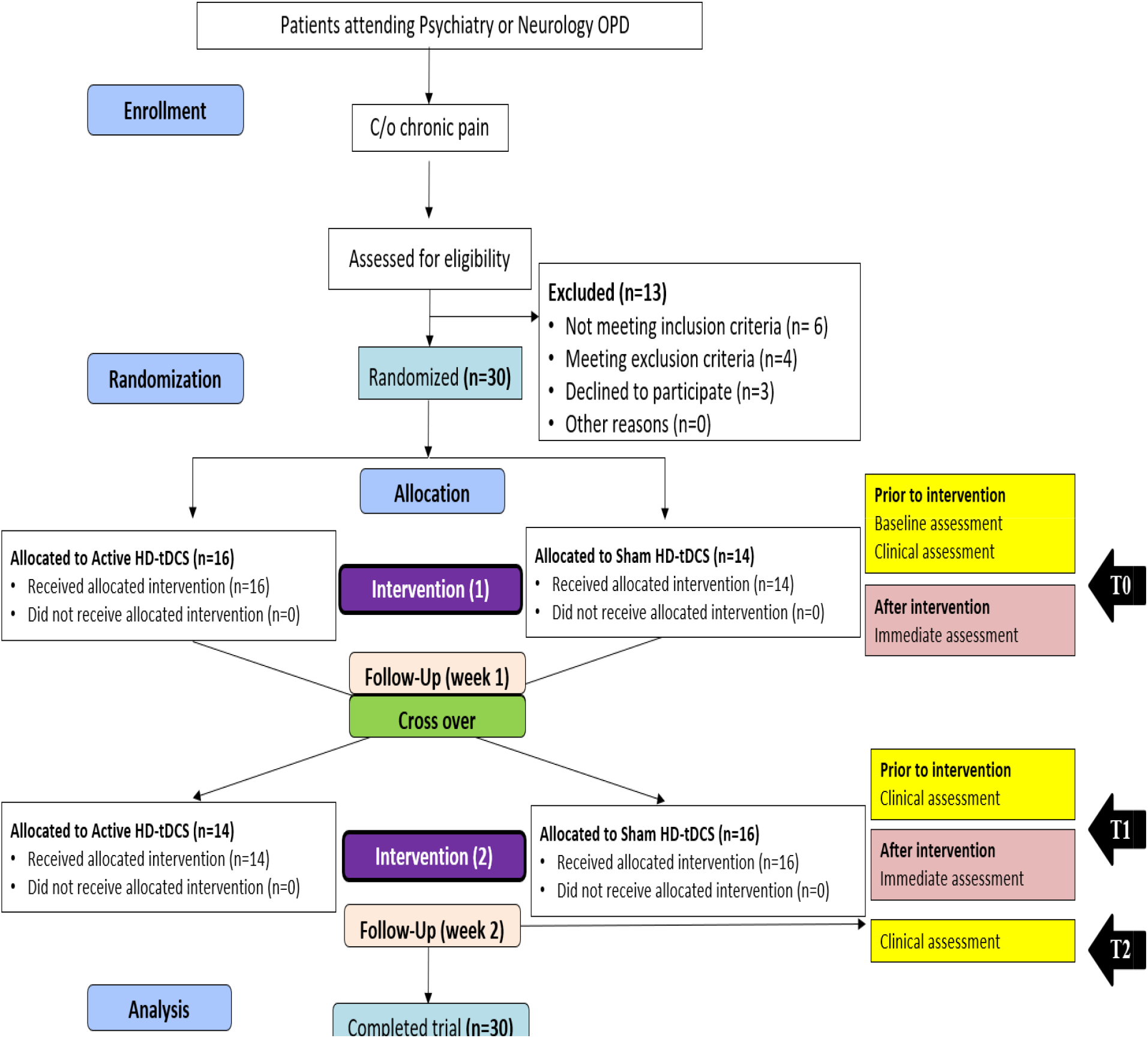
Participant flow diagram.

### 2.5 Intervention

One block comprised of two sessions of 20 minutes each of same type (either active-active or sham-sham) HD-tDCS, with 30 min interval delivered by standard HD-tDCS equipment. In the active block, a 2mA current was delivered after a ramp up time of 30 seconds. In case of sham, there was a brief period of stimulation in which patient would have a sensation of current flowing, but no actual effect would occur. Intervention consisted of one block of HD-tDCS crossed over from active to sham and vice versa after a wash out period of one week, thereby every patient received both active and sham stimulation at the end of two weeks. Motor hotspot was detected using the visual method by evoking contraction of the Abductor pollicis brevis muscle of the right hand using single pulse transcranial magnetic stimulus.

### 2.6 Randomisation

A consecutive sampling technique and simple randomisation was utilized. Eligible patients were randomized in 1:1 allocation ratio by computer assigned random allocation to one of two treatment arms: active or sham HD-tDCS and concealed in sealed envelopes. The random allocation sequence was generated by one of the authors, while enrolling of participants and assigning them to the treatment sequence was done by another author. Patients were blinded to the treatment sequence. In view of the reduction in sample size, 16 patients were allocated to active followed by sham treatment arm and 14 patients were allocated to sham followed by active treatment arm.

### 2.7 Analysis

Descriptive analysis in terms of frequencies, means & standard deviations, Chi Square tests for ordinal data and t tests for normally distributed data were employed. Repeated measures analysis of variance (ANOVA) to test the hypothesis regarding the effect of HD-tDCS on the pain scores with treatment order (active-sham versus sham-active), treatment condition (active versus sham), and time (baseline, immediately after stimulation, and at follow-ups) as the independent variables was also applied. Analysis of Covariance (ANCOVA) to test interaction of order and treatment group was applied. Statistical analysis was done using SPSS software version 21. CONSORT guidelines(Dwan et al., 2019) are presented in supplementary Table 1.

## 3. Results

### 3.1 Socio demographic and clinical profile

More than 70% patients were female and had moderate to severe somatic symptoms. Comorbid physical conditions were seen in one fourth patients, commonest being hypertension followed by diabetes and hypothyroidism.

Patients were on various medication like Amitriptyline (25-50 mg per day), Duloxetine (30-120 mg per day), Escitalopram (10-20 mg per day), Gabapentin (40 mg per day), Imipramine (75-200 mg per day), Nortriptyline (10 mg per day), Pregabalin (75-150 mg per day), Propranolol (20-40 mg per day), Sertraline (150 mg per day) and Venlafaxine (150 mg per day). 6 patients were on a combination of the 2 of the above mentioned medications.

Mean baseline pain score on 10 point NRS was 7.93 (1.85), which was almost similar when rated on BPI. Moderate levels of anxiety and depressive symptoms were observed at baseline.

### 3.2 Comparison of baseline characteristics between groups

Groups A (active stimulation followed by sham, n=16) and B (sham stimulation followed by active, n=14) were comparable on all domains of the socio-demographic and clinical profile, various pain scores and other scores of pain related interference, anxiety, depressive symptoms, burden, disability and cognitive functions with no statistically significant difference. This is summarized in Table 1.

**Table 1.** Socio-demographic and baseline clinical profile.

| **Parameters** | **Total patients**<br>**(N= 30)**<br>**Frequency (%) or**<br>**Mean (SD) [Range]** | **Group A**<br>**(active followed**<br>**by sham)**<br>**(N=16)**<br>**Frequency (%)**<br>**or Mean (SD)** | **Group B**<br>**(sham followed**<br>**by active)**<br>**(N=14)**<br>**Frequency (%)**<br>**or Mean (SD)** |
| --- | --- | --- | --- |
| Age (years) | 39.90 ( 9.88) [22-59] | 40.44 (8.93) | 39.29 (11.18) |
| Education (years) | 10.83 ( 6.41) [ 0-23] | 11.00 (5.81) | 10.64 ( 7.26) |
| Gender |  |  |  |
| Male | 8 (26.7%) | 3 (18.7%) | 5 (35.7%) |
| Female | 22 (73.3%) | 13 (81.3%) | 9 (64.3%) |
| Duration of illness (years) | 8.57 ( 6.17) [ 1-25] | 8.44 (5.27) | 8.71 ( 7.28) |
| DSM-5 severity of somatic<br>symptoms |  |  |  |
| Mild to moderate | 20 (66.7%) | 11 (68.8%) | 9 (64.3%) |
| Severe | 10 (33.3%) | 5 (31.2%) | 5 (35.7%) |
| PHQ-15 | 12.90 ( 6.81) [4-28] | 12.75 ( 7.36) | 13.07 ( 6.40) |
| Comorbid physical<br>diagnosis | 8 (26.7%) | 4 (25.0%) | 4 (28.6%) |
| Comorbid substance use<br>(alcohol) | 3 (10%) | 0 ( 0%) | 3 (21.5%) |
| NRS | 7.93 ( 1.86) [5-10] | 7.50 ( 1.97) | 8.42 ( 1.65) |
| BPI-H (average pain) | 7.33 ( 1.88) [4-10] | 7.00 ( 1.75) | 7.71 ( 2.02) |
| BPI-H (interference) | 6.56 ( 1.99) [1.7-9.6] | 6.10 ( 2.22) | 7.09 ( 1.63) |
| GAD-7 | 12.40 ( 5.62) [ 0-21] | 12.12 ( 6.30) | 12.71 ( 4.97) |
| PHQ- 9 | 13.66 ( 5.62) [ 1-27] | 13.12 ( 6.37) | 14.28 ( 4.76) |
| SSS- 8 | 17.36 ( 8.31) [ 4-31] | 17.56 ( 8.79) | 17.14 ( 8.06) |
| PDI- 7 | 39.73 (15.29) [ 6-68] | 38.31 (11.92) | 41.35 (18.78) |
| MMSE | 29.73 ( 0.69) [28-30] | 29.62 ( 0.81) | 29.85 ( 0.53) |
| CGI-S | 4.06 ( 0.78) [ 3- 6] | 4.06 ( 0.77) | 4.07 ( 0.83) |

SD: Standard deviation, %: Percentage, N: Total number, PHQ-15: Patient Health Questionnaire 15, NRS: Numerical Rating Scale, BPI-H: Brief Pain Inventory-Hindi version, GAD-7: Generalized Anxiety Disorder 7 Scale, PHQ-9: Patient Health Questionnaire 9, SSS-8: Somatic Symptom Scale–8, PDI-7: Pain Disability Index 7, MMSE: Mini Mental State Assessment CGI-S: Clinical Global Impression- severity

### 3.3 Results comparing overall active versus sham stimulation

Outcome of active stimulation was compared to sham stimulation (n=30). This is summarized in Table 2. Outcome of pain scores were statistically significant between active and sham groups on NRS and BPI. Outcome of interference due to pain, anxiety, depressive symptoms, burden and disability due to pain showed statistically significant difference between active and sham groups. But there was no difference on cognitive function scores between the groups.

**Table 2.** Outcome of pain and other associated scores in active vs sham groups.

| Outcome variables | Paired differences |  |  |  | t-<br>value | p-value |
| --- | --- | --- | --- | --- | --- | --- |
|  | Mean<br>(SD) | SE of<br>mean | 95% confidence<br>interval of the<br>difference |  |  |  |
|  |  |  | lower | upper |  |  |
| NRS | -0.80 (0.99) | 0.182 | -1.172 | -0.428 | -4.397 | <**0.01** |
| BPI-H (average pain) | -0.56 (0.93) | 0.171 | -9.159 | -0.217 | -3.319 | <**0.01** |
| BPI-H (interference) | -0.56 (0.77) | 0.135 | -0.845 | -0.288 | -4.171 | <**0.01** |
| GAD-7 | -1.53 (2.43) | 0.444 | -2.441 | -0.625 | -3.454 | <**0.01** |
| PHQ- 9 | -1.66 (2.88) | 0.526 | -2.742 | -0.591 | -3.169 | <**0.01** |
| SSS- 8 | -2.00 (3.02) | 0.553 | -3.131 | -0.869 | -3.617 | <**0.01** |
| PDI- 7 | -3.03 (4.15) | 0.758 | -4.585 | -1.481 | -3.997 | <**0.01** |
| MMSE | 0 | 0 | - | - | - | - |

SD: Standard deviation, SE: Standard error, df: degree of freedom, NRS: Numerical Rating Scale, BPI-H: Brief Pain Inventory-Hindi version, GAD-7: Generalized Anxiety Disorder 7 Scale, PHQ-9: Patient Health Questionnaire 9, SSS-8: Somatic Symptom Scale–8, PDI-7: Pain Disability Index 7, MMSE: Mini Mental State Assessment

### 3.4 Side effect profile

Table 3 summarises the side effects seen in both groups. Statistically significant difference was seen between active and sham stimulation in burning and pain, but not in itching. No severe side effects like insomnia, acute mood changes or changes in visual perception were noted.

**Table 3.** Frequency and types of side effects experienced after active vs sham stimulation.

| Side effects checklist | Active<br>stimulation<br>(N=30)<br>Frequency (%) | Sham<br>stimulation<br>(N=30)<br>Frequency (%) | Chi- square value<br>(p-value) |
| --- | --- | --- | --- |
| Numbness at stimulation site | 0 (0%) | 0 (0%) | - |
| Redness at stimulation site | 0 (0%) | 0 (0%) | - |
| Itching at stimulation site | 9 (30.0%) | 6 (20.0%) | 0.813 (0.371) |
| Burning at stimulation site | 25 (83.3%) | 12 (40.0%) | **11.915 (<0.01)** |
| Pain at stimulation site | 16 (53.3%) | 7 (23.3%) | **5.711 (0.016)** |
| Nausea | 0 (0%) | 0 (0%) | - |
| Fatigue | 0 (0%) | 0 (0%) | - |
| Nervousness | 0 (0%) | 0 (0%) | - |
| Insomnia | 0 (0%) | 0 (0%) | - |
| Headache | 0 (0%) | 0 (0%) | - |
| Difficulty in concentrating | 0 (0%) | 0 (0%) | - |
| Acute mood changes | 0 (0%) | 0 (0%) | - |
| Changes in visual perception | 0 (0%) | 0 (0%) | - |

?: Percentage, N: Total number

### 3.5 Results based on entry into active versus sham stimulation

One way ANCOVA was run to look for interaction of the order of intervention (active followed by sham and sham followed by active) on the pain outcome on NRS. Error of variance across the groups was equal as per Levene’s test of equality, and so the data met the homogeneity of variances assumption. Order of intervention by treatment group interaction is not statistically significant [F (1, 55)= 2.136 (p = 0.150)].

As shown in table 4, when entry status of active and sham stimulation was considered, there was no statistically significant difference between baseline (T0) and week 2 (T2) scores in both groups A and B on NRS. But there was statistically significant difference between baseline (T0) and week one (T1) scores in group A only and between week one (T1) and week two (T2) in group B only. The same was observed with respect to all other outcome measures on RM ANOVA.

**Table 4.** Outcome of pain on NRS as per entry order of groups A and B.

| Group | Mean (SD) |  |  | RM ANOVA |  | T0 vs T2 |  | T0 vs T1 |  | T1 vs T2 |  |
| --- | --- | --- | --- | --- | --- | --- | --- | --- | --- | --- | --- |
| | T0 | T1 | T2 | p | $\eta_p^2$ | MD (SE) | p | MD (SE) | p | MD (SE) | p |
| A (N=16) | 7.50<br>(1.97) | 4.18<br>(2.29) | 4.18<br>(2.29) | <0.01 | 0.817 | 3.313<br>(0.405) | <0.01 | 3.313<br>(0.405) | <0.01 | 0 (0) | - |
| B (N=14) | 8.42<br>(1.65) | 7.92<br>(2.02) | 6.21<br>(2.22) | <0.01 | 0.726 | 2.214<br>(0.334) | <0.01 | 0.500<br>(0.292) | 0.331 | 1.714<br>(0.194) | <0.01 |

SD: Standard deviation, N: Total number, p: p-value, $\eta_p^2$ : partial eta-squared, MD: Mean difference, SE: Standard error, T0: baseline value, T1: week 1 value, T2: week 2 value, NRS: Numerical Rating Scale

### 3.6 Global improvement

Clinician rated global improvement on CGI-efficacy index at week one showed statistically significant difference between active and sham stimulation, but no statistically significant difference at week two. Similarly, patient rated global improvement on PGIC at week one showed statistically significant difference between active and sham stimulation, but no statistically significant difference at week two.

## 4. Discussion

To our knowledge, this is one of the first studies of HD-tDCS in SSD-P. Patients were on stable dose of pharmacological treatment for at least 3 months during recruitment for HD-tDCS as add on therapy. Despite adequate treatment, only three patients refused to participate in the study which shows the acceptability of the intervention among patients of SSD-P. Though our primary focus was on effect on pain, a host of secondary outcomes like interference due to pain, burden due to somatic symptoms, depressive symptoms, anxiety symptoms, disability due to pain and cognitive features were also assessed using standard rating scales which gives a holistic approach to pain.

Cross over study design was employed, which is recommended and commonly utilized in trials on pain treatment and known to reduce between-patient variability(Wellek and Blettner, 2012). Moreover, permuted block randomization was done to rule out any potential bias due to order of active or sham stimulation(Biabani et al., 2018). In addition, one way ANCOVA with order as covariate showed no interaction of order and group. Blinding was ensured as patients were unable to discern above chance (p=0.209) whether they had received active or sham stimulation. The ramp up and down at the beginning and end of sham stimulation simulates active stimulation, making the patient perceive sensation of current flowing(Kessler et al., 2012). There was no statistically significant difference between patients blinded to active and sham groups at entry on any of the sociodemographic or clinical parameters, eliminating any possible confounder bias between the groups at their entry into the study(Biabani et al., 2018).

Differences between the groups as per entry status at T0 VS T1 in first active group and T1 VS T2 in first sham group shows that only active intervention had any impact on reduction in pain scores when given first or second, irrespective of order. Sham stimulation given either as first or second session showed no significant differences on the outcome parameters. This could also mean that in the group receiving active stimulation, there was no carry over effect when sham stimulation was provided.

Apart from use of HD-tDCS, focal stimulation was also ensured by single pulse TMS aided marking of the left motor cortex. When 2 within session repeated HD-tDCS was delivered, we observed sustained effects that lasted for one week. This was demonstrated in other studies which showed that sustained effects could be observed only in short interval within session repeated tDCS, but not with longer intervals (beyond two to three hours)(Monte-Silva et al., 2010) or use of higher amplitudes (3mA)(Agboada et al., 2020). The sustained effects are said to be due to stimulation-timing dependent plasticity regulation in the cortex(Brunoni et al., 2012). We observed high Cohen’s d of 0.85 for reduction in pain and other associated variables. This shows that our protocol is effective.

Patients experienced short lasting side effects of mild to moderate intensity, similar to other studies on tDCS(Matsumoto and Ugawa, 2017). No cognitive or other serious side effects were seen. Specialized electrodes used in HD-tDCS ensures precise control of contact conditions, unlike conventional tDCS which shows that HD-tDCS is well tolerated(Villamar et al., 2013).

To our knowledge, this is one of the first studies to study the effect of HD-tDCS on pain in SSD-P. Simple protocol of two repeat sessions of HD-tDCS was administered. Holistic approach by standard assessment using appropriate rating scales was done on pain and its associated variables. The study was feasible as it was completed in a one year period. There was no drop out and hence the study can be considered acceptable. Any potential bias due to blinding, allocation, baseline patient characteristics, order effect or carry over effect was ruled out.

Use of a cross over study design although recommended for trials of pain treatment, may add to bias. Small sample size which was further reduced due to the COVID-19 restrictions and lockdown and single blinding are limitations of this study.

## 5. Conclusion

The study shows that two within session repeated HD-tDCS is effective and tolerable in reduction of pain and other associated variables in SSD-P and effects are sustained for a week. There is a need for studies using more than one type of single session protocols and comparison between them to come up with an ideal protocol. Formulation of maintenance protocols that will carry forward the plasticity for longer lasting periods leading to better quality of life will benefit the patients. More studies evaluating parameters other than pain are required to get a holistic idea of the effectiveness of neuromodulation.

## Data Availability

All data produced in the present study are available upon reasonable request to the authors

## Acknowledgements

Nil

## References

Agboada, D., Mosayebi-Samani, M., Kuo, M.-F., Nitsche, M.A., 2020. Induction of long-term potentiation-like plasticity in the primary motor cortex with repeated anodal transcranial direct current stimulation – Better effects with intensified protocols? Brain Stimulation 13, 987–997. https://doi.org/10.1016/j.brs.2020.04.009

American Psychiatric Association, 2013. Diagnostic and Statistical Manual of Mental Disorders (DSM-5®), 5th ed. American Psychiatric Publishing, Arlington, VA.

Bastani, A., Jaberzadeh, S., 2014. Within-session repeated a-tDCS: The effects of repetition rate and inter-stimulus interval on corticospinal excitability and motor performance. Clin Neurophysiol 125, 1809–1818. https://doi.org/10.1016/j.clinph.2014.01.010

Biabani, M., Farrell, M., Zoghi, M., Egan, G., Jaberzadeh, S., 2018. Crossover design in transcranial direct current stimulation studies on motor learning: potential pitfalls and difficulties in interpretation of findings. Rev Neurosci 29, 463–473. https://doi.org/10.1515/revneuro-2017-0056

Bolognini, N., Olgiati, E., Maravita, A., Ferraro, F., Fregni, F., 2013. Motor and parietal cortex stimulation for phantom limb pain and sensations. Pain 154, 1274–1280. https://doi.org/10.1016/j.pain.2013.03.040

Brunoni, A.R., Nitsche, M.A., Bolognini, N., Bikson, M., Wagner, T., Merabet, L., Edwards, D.J., Valero-Cabre, A., Rotenberg, A., Pascual-Leone, A., Ferrucci, R., Priori, A., Boggio, P., Fregni, F., 2012. Clinical Research with Transcranial Direct Current Stimulation (tDCS): Challenges and Future Directions. Brain Stimul 5, 175–195. https://doi.org/10.1016/j.brs.2011.03.002

Dimsdale, J.E., Creed, F., Escobar, J., Sharpe, M., Wulsin, L., Barsky, A., Lee, S., Irwin, M.R., Levenson, J., 2013. Somatic symptom disorder: an important change in DSM. J Psychosom Res 75, 223–228. https://doi.org/10.1016/j.jpsychores.2013.06.033

Donnell, A., D Nascimento, T., Lawrence, M., Gupta, V., Zieba, T., Truong, D.Q., Bikson, M., Datta, A., Bellile, E., DaSilva, A.F., 2015. High-Definition and Non-invasive Brain Modulation of Pain and Motor Dysfunction in Chronic TMD. Brain Stimul 8, 1085–1092. https://doi.org/10.1016/j.brs.2015.06.008

Dwan, K., Li, T., Altman, D.G., Elbourne, D., 2019. CONSORT 2010 statement: extension to randomised crossover trials. BMJ 366, l4378. https://doi.org/10.1136/bmj.l4378

Fenton, B.W., Palmieri, P.A., Boggio, P., Fanning, J., Fregni, F., 2009. A preliminary study of transcranial direct current stimulation for the treatment of refractory chronic pelvic pain. Brain Stimul 2, 103–107. https://doi.org/10.1016/j.brs.2008.09.009

Henningsen, P., 2018. Management of somatic symptom disorder. Dialogues Clin Neurosci 20, 23–31.

Janice Jimenez-Torres, G., Weinstein, B.L., Walker, C.R., Christopher Fowler, J., Ashford, P., Borckardt, J.J., Madan, A., 2017. A study protocol for a single-blind, randomized controlled trial of adjunctive transcranial direct current stimulation (tDCS) for chronic pain among patients receiving specialized, inpatient multimodal pain management. Contemp Clin Trials 54, 36–47. https://doi.org/10.1016/j.cct.2016.12.024

Katz, J., Rosenbloom, B.N., Fashler, S., 2015. Chronic Pain, Psychopathology, and DSM-5 Somatic Symptom Disorder. Can J Psychiatry 60, 160–167. https://doi.org/10.1177/070674371506000402

Kessler, S.K., Turkeltaub, P.E., Benson, J.G., Hamilton, R.H., 2012. Differences in the Experience of Active and Sham Transcranial Direct Current Stimulation. Brain Stimul 5, 155–162. https://doi.org/10.1016/j.brs.2011.02.007

Kleinstäuber, M., Witthöft, M., Steffanowski, A., van Marwijk, H., Hiller, W., Lambert, M.J., 2014. Pharmacological interventions for somatoform disorders in adults. Cochrane Database Syst Rev CD010628. https://doi.org/10.1002/14651858.CD010628.pub2

Lefaucheur, J.-P., 2016. A comprehensive database of published tDCS clinical trials (2005– 2016). Neurophysiologie Clinique/Clinical Neurophysiology 46, 319–398. https://doi.org/10.1016/j.neucli.2016.10.002

Lefaucheur, J.-P., Antal, A., Ayache, S.S., Benninger, D.H., Brunelin, J., Cogiamanian, F., Cotelli, M., De Ridder, D., Ferrucci, R., Langguth, B., Marangolo, P., Mylius, V., Nitsche, M.A., Padberg, F., Palm, U., Poulet, E., Priori, A., Rossi, S., Schecklmann, M., Vanneste, S., Ziemann, U., Garcia-Larrea, L., Paulus, W., 2017. Evidence-based guidelines on the therapeutic use of transcranial direct current stimulation (tDCS). Clin Neurophysiol 128, 56–92. https://doi.org/10.1016/j.clinph.2016.10.087

Matsumoto, H., Ugawa, Y., 2017. Adverse events of tDCS and tACS: A review. Clinical Neurophysiology Practice 2, 19–25. https://doi.org/10.1016/j.cnp.2016.12.003

Mendonca, M.E., Santana, M.B., Baptista, A.F., Datta, A., Bikson, M., Fregni, F., Araujo, C.P., 2011. Transcranial DC Stimulation in Fibromyalgia: Optimized Cortical Target Supported by High-Resolution Computational Models. The Journal of Pain 12, 610– 617. https://doi.org/10.1016/j.jpain.2010.12.015

Monte-Silva, K., Kuo, M.-F., Liebetanz, D., Paulus, W., Nitsche, M.A., 2010. Shaping the optimal repetition interval for cathodal transcranial direct current stimulation (tDCS). J Neurophysiol 103, 1735–1740. https://doi.org/10.1152/jn.00924.2009

Ngernyam, N., Jensen, M.P., Arayawichanon, P., Auvichayapat, N., Tiamkao, S., Janjarasjitt, S., Punjaruk, W., Amatachaya, A., Aree-uea, B., Auvichayapat, P., 2015. The effects of transcranial direct current stimulation in patients with neuropathic pain from spinal cord injury. Clin Neurophysiol 126, 382–390. https://doi.org/10.1016/j.clinph.2014.05.034

Raja, S.N., Carr, D.B., Cohen, M., Finnerup, N.B., Flor, H., Gibson, S., Keefe, F.J., Mogil, J.S., Ringkamp, M., Sluka, K.A., Song, X.-J., Stevens, B., Sullivan, M.D., Tutelman, P.R., Ushida, T., Vader, K., 2020. The revised International Association for the Study of Pain definition of pain: concepts, challenges, and compromises. Pain 161, 1976– 1982. https://doi.org/10.1097/j.pain.0000000000001939

Tortella, G., Casati, R., Aparicio, L.V.M., Mantovani, A., Senço, N., D’Urso, G., Brunelin, J., Guarienti, F., Selingardi, P.M.L., Muszkat, D., Junior, B. de S.P., Valiengo, L., Moffa, A.H., Simis, M., Borrione, L., Brunoni, A.R., 2015. Transcranial direct current stimulation in psychiatric disorders. World J Psychiatry 5, 88–102. https://doi.org/10.5498/wjp.v5.i1.88

van Dessel, N., den Boeft, M., van der Wouden, J.C., Kleinstäuber, M., Leone, S.S., Terluin, B., Numans, M.E., van der Horst, H.E., van Marwijk, H., 2014. Non-pharmacological interventions for somatoform disorders and medically unexplained physical symptoms (MUPS) in adults. Cochrane Database Syst Rev CD011142. https://doi.org/10.1002/14651858.CD011142.pub2

Villamar, M.F., Volz, M.S., Bikson, M., Datta, A., DaSilva, A.F., Fregni, F., 2013. Technique and Considerations in the Use of 4×1 Ring High-definition Transcranial Direct Current Stimulation (HD-tDCS). JoVE 50309. https://doi.org/10.3791/50309

Wellek, S., Blettner, M., 2012. On the Proper Use of the Crossover Design in Clinical Trials. Dtsch Arztebl Int 109, 276–281. https://doi.org/10.3238/arztebl.2012.0276

